# A Highly Accurate Ensemble Classifier for the Molecular Diagnosis of ASD at Ages 1 to 4 Years

**DOI:** 10.1101/2021.07.08.21260225

**Authors:** Bokan Bao, Vahid H. Gazestani, Yaqiong Xiao, Raphael Kim, Austin W.T. Chiang, Srinivasa Nalabolu, Karen Pierce, Kimberly Robasky, Nathan E. Lewis, Eric Courchesne

## Abstract

**Importance:** ASD diagnosis remains behavior-based and the median age of the first diagnosis remains unchanged at ∼52 months, which is nearly 5 years after its first trimester origin. Long delays between ASD’s prenatal onset and eventual diagnosis likely is a missed opportunity. However, accurate and clinically-translatable early-age diagnostic methods do not exist due to ASD genetic and clinical heterogeneity. There is a need for early-age diagnostic biomarkers of ASD that is robust against its heterogeneity.

**Objective:** To develop a single blood-based molecular classifier that accurately diagnoses ASD at the age of first symptoms.

**Design, Setting, and Participants:** N=264 ASD, typically developing (TD), and language delayed (LD) toddlers with their clinical, diagnostic, and leukocyte RNA data collected. Datasets included Discovery (n=175 ASD, TD subjects), Longitudinal (n=33 ASD, TD subjects), and Replication (n=89 ASD, TD, LD subjects). We developed an ensemble of ASD classifiers by testing 42,840 models composed of 3,570 feature selection sets and 12 classification methods. Models were trained on the Discovery dataset with 5-fold cross validation. Results were used to construct a Bayesian model averaging-based (BMA) ensemble classifier model that was tested in Discovery and Replication datasets. Data were collected from 2007 to 2012 and analyzed from August 2019 to April 2021.

**Main Outcomes and Measures:** Primary outcomes were (1) comparisons of the performance of 42,840 classifier models in correctly identifying ASD vs TD and LD in Discovery and Replication datasets; and (2) performance of the ensemble model composed of 1,076 models and weighted by Bayesian model averaging technique.

**Results:** Of 42,840 models trained in the Discovery dataset, 1,076 averaged AUC-ROC>0.8. These 1,076 models used 191 different feature routes and 2,764 gene features. Using weighted BMA of these features and routes, an ensemble classifier model was constructed which demonstrated excellent performance in Discovery and Replication datasets with ASD classification AUC-ROC scores of 84% to 88%. ASD classification accuracy was comparable against LD and TD subjects and in the Longitudinal dataset. ASD toddlers with ensemble scores above and below the ASD ensemble mean had similar diagnostic and psychometric scores, but those below the ASD ensemble mean had more prenatal risk events than TD toddlers. Ensemble features include genes with immune/inflammation, response to cytokines, transcriptional regulation, mitotic cell cycle, and PI3K-AKT, RAS, and Wnt signaling pathways.

**Conclusions and Relevance:** An ensemble ASD molecular classifier has high and replicable accuracy across the spectrum of ASD clinical characteristics and across toddlers aged 1 to 4 years, which has potential for clinical translation.

**Key Points:** *Question:* Since ASD is genetically and clinical heterogeneous, can a single blood-based molecular classifier accurately diagnose ASD at the age of first symptoms?

*Findings:* To address heterogeneity, we developed an ASD classifier method testing 42,840 models. An ensemble of 1,076 models using 191 different feature routes and 2,764 gene features, weighted by Bayesian model averaging, demonstrated excellent performance in Discovery and Replication datasets producing ASD classification with the area under the receiver operating characteristic curve (AUC-ROC) scores of 84% to 88%. Features include genes with immune/inflammation, response to cytokines, transcriptional regulation, mitotic cell cycle, and PI3K-AKT, RAS and Wnt signaling pathways.

*Meaning:* An ensemble gene expression ASD classifier has high accuracy across the spectrum of ASD clinical characteristics and across toddlers aged 1 to 4 years.

## INTRODUCTION

ASD is a prenatal^1–16^, highly heritable disorder^17^ that significantly impacts a child’s ability to perceive and react to social information^18–20^. Despite this prenatal and strongly genetic beginning, robust and replicable early-age biological ASD diagnostic markers useful at the individual level have not been found. Across the past two decades, ASD diagnosis remains behavior-based and the median age of the first diagnosis remains unchanged at ∼52 months^21–24^, which is nearly 5 years after its first trimester origin. The long delay between ASD’s prenatal onset and eventual diagnosis is a missed opportunity. Moreover, the heterogeneity of ASD genetics and clinical characteristics has imposed barriers to the identification of early-age diagnostic biomarkers that accurately diagnose the broad majority of those with this heterogeneous disorder^25^. Thus, there is a need for early-age diagnostic biomarkers of ASD that can robustly fight against this heterogeneity.

Since ASD has a heritability of 81%^17^, initial attempts focused on genetics as a way towards the discovery of clinically useful biomarkers for precision medicine as well as causal explanations for ASD pathogenesis. While syndromic risk mutations have been described for >200 genes in ASD^15,26^, each occurs only rarely in ASD. For 80% to 90% of patients, such mutations are not found. Thus, an estimated 80% of ASD individuals are still considered ‘idiopathic’, wherein little is known about the specific genes and/or environmental factors causing their disorder. In this idiopathic majority of ASD, the risk is likely associated with a large number of inherited common and rare risk variants in each individual child. Studies of polygenic ASD risk found that the combined effect of genetic risk variants in case-control studies accounts for less than 7.5% of the risk variance^27^; genetic ASD risk scores substantially overlap with controls^26^; and polygenic risk scores are not clinically diagnostic or prognostic at the individual level nor explanatory for the majority of ASD^28^. Thus, DNA-based mutations or polygenic risk scores may not be useful for the many idiopathic ASD subjects at the translational level.

RNA biomarkers have been sought using blood gene expression in more than 35 ASD studies since 2006, but the vast majority have been underpowered, older-aged, clinically heterogeneous, and absent replication datasets^29–39^. Some early genetics researchers rejected blood-based biomarkers believing that ASD-relevant dysregulated gene expression must be restricted to the brain. Recent ASD genetics have turned this view on its head: The earliest prenatal drivers of deviant ASD development are, in fact, broadly expressed regulatory genes that are active in blood leukocytes as well as other organs including the prenatal brain^1–3,29,30,33,34,39–41^. Broadly expressed genes that constitute the majority of ASD risk genes are upregulated in the early prenatal life and impact multiple stages of prenatal brain development from the first- and second-trimester proliferation and neurogenesis to neurite outgrowth and synaptogenesis in the third trimester. They do so by disrupting gene expression in signaling pathways such as PI3K-AKT, RAS-ERK, and Wnt signaling pathways^42^, which, in turn, disrupt prenatal functions.

In idiopathic ASD toddlers aged 1-4 year olds, leukocyte gene expression in these pathways is significantly dysregulated. At the group level, the degree of dysregulation is correlated with ASD social symptom severity^43^. Broadly expressed genes in leukocytes from ASD toddlers are also associated with hypoactive brain responses to language, dysregulation of ASD and language relevant genes, and poor language outcomes^30^. Thus, leukocyte gene expression holds the potential for the objective identification of biological subtypes of ASD. In analyses of leukocyte gene co-expression, ASD-associated module eigengene values were significantly correlated with abnormal early brain growth and enriched in genes related to cell cycle, translation, and immune networks and pathways. These same gene sets are very accurate classifiers of ASD vs control toddlers^30^. Meta-analyse or mega-analyse of the wider ASD blood gene expression literature and individual studies^29–39^ find that there appears to be dysregulated gene expression in PI3K-AKT-mTOR, RAS signaling pathways, increased ribosomal translation signal, and dysregulation of genes involved in cell cycle, inflammation-related processes, interferon signaling, and the KEGG natural killer cytotoxicity pathway.

Leukocyte gene expression patterns offer a non-invasive and clinically practicable avenue for understanding aspects of ASD cell biology, including those that could be ASD-relevant, ASD-specific, robust, and diagnostic or prognostic. However, if the clinical translational potential of leukocyte transcriptomics is to lead to robust and rigorous classifiers, then high standards for verifying such classifiers should be implemented.

Here we developed, operationalized, and tested a rigorous analytic pipeline to identify molecular diagnostic classifiers for ASD using leukocyte gene expression. Using this platform on leukocyte transcriptomic data from male ASD toddlers at ages 1-4 years old, typically developing (TD), and language delayed (LD) subjects, we systematically analyzed the classification performance of 42,840 different models composed of 3,570 different feature selection sets and 12 commonly-used classification methods (**Figure 1A**). In this way, we developed a highly accurate ensemble diagnostic classifier of male ASD toddlers.

**Figure 1.**
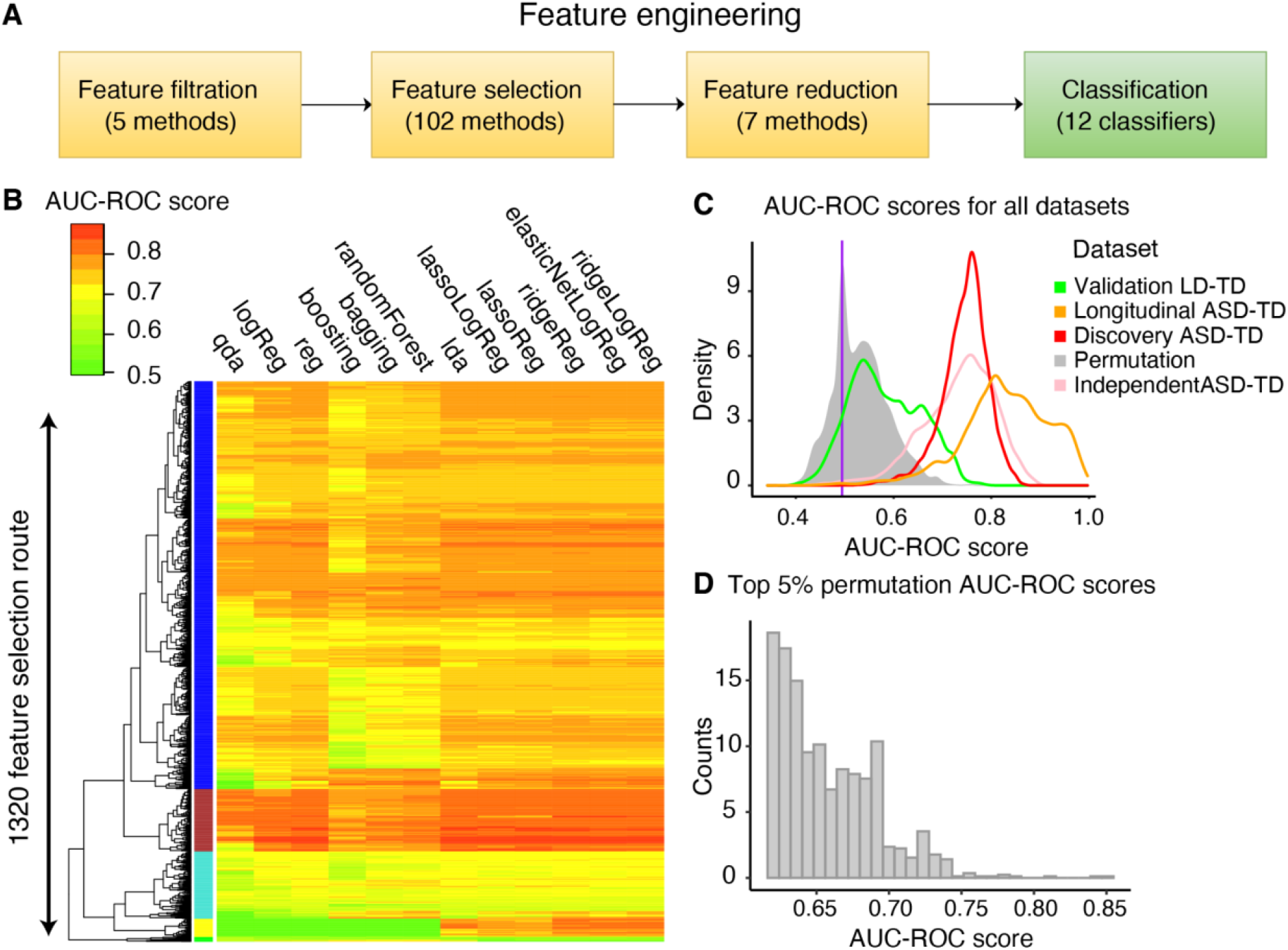
A classification platform was developed to robustly identify the biomarkers for the early diagnosis of ASD. **A**. Our platform performs 3570 feature engineering methods, followed by 12 different classification approaches on the input datasets. **B**. AUC-ROC values were computed for each feature selection route and classification method on the Discovery dataset. The AUC-ROC values are based on the average model performance across 5 iterations, with 20% of samples being held out each time. **C**. 15,840 models were further tested on an independent Replication dataset and Longitudinal samples from the Discovery dataset, and on average performed well. Evaluation of the models on a dataset of language delay subjects showed the specificity of the signals to ASD subjects. Permuting the sample labels (i.e., ASD and TD) further supported the validity of the signal. **D**. Top 5% AUC-ROC score of permutation data.

## RESULTS

### Development of a robust transcriptomic classifier platform with diverse feature engineering and classification methods

To identify potential blood transcriptome biomarkers in our Discovery sample of 175 ASD and TD toddlers (Table 1), we developed a platform that robustly reveals the classification power of blood transcriptome by systematically exploring the performance of 42,840 possible models composed of 3570 different feature selection routes followed by 12 classification methods (see **Methods, Figure 1A**). As shown in **Figure 1A**, the platform started with removing genes with low variation across samples. Next, features that differentiate between ASD and TD subjects at expression or co-expression levels were selected using a suite of 102 feature selection methods. Third, to avoid overfitting, we reduced the number of features by collapsing expression data from the correlating genes. Finally, we trained 12 different classifiers based on each of the selected feature sets. To robustly evaluate the performance of each of the 3570 feature selection routes and the 12 classification methods, we iterated the process 5 times while holding out 20% of samples and using the remaining 80% of samples for hyper-parameter selection, feature selection, and classifier training. Thus, each of the 42,840 models started with a “route” that consisted of 1 filtration method, 1 selection method, 1 reduction method, and ended with 1 classification method. The platform reports the average performance of each of the 42,840 models across the 5 held-out folds as measured by area under the receiver operating characteristic curve (AUC-ROC) and area under the precision-recall curve (AUC-PR).

**Table 1.** Summary of the Subjects.

| Dateset | ASD | TD | LD |
| --- | --- | --- | --- |
| Discovery | 93 | 82 | 0 |
| Independent Replication | 34 | 31 | 24 |
| Longitudinal | 18 | 15 | 0 |

ASD: autism spectrum disorder; TD: typical development; LD: language delayed

### Diverse pipelines successfully classify ASD vs TD

Since the feature selection methods depended on the characteristics of training transcriptome datasets, some routes were not able to generate features in all five iterations of the platform. Accordingly, the platform successfully classified the data in 15,840 out of 42,840 different ways, including 1320 different routes out of 3570 for feature selection and 12 different classification methods (Supplementary Table 1). From 15,840 trained models, 1822 (11.5%) models showed classification AUC-ROC of above 0.8 with the max AUC-ROC of 0.856 (**Figure 1B**, Supplementary Figure 3). Moreover, 1508 of the 1822 models also exhibited an AUC-PR of above 0.8.

These 1822 high-performing models performed well due to their feature routes and were robust to variations in the data or the model. For example, we observed a subset of 175 feature routes (colored with a brown band in **Figure 1B)** that performed consistently well across different classifiers with a mean AUC-ROC score of 0.810. Additionally, these 1822 high-performing models worked similarly well across all five held-out datasets with a mean range of 0.13 and variance of 0.02 (Supplementary Figure 4). Furthermore, different models that largely overlapped in their feature selection routes also worked well across different classifier methods (Supplementary Figure 3).

To further verify that the performance of these 1822 models was not due to chance alone, we generated five separate randomized datasets by shuffling the sample labels (i.e., ASD or TD) from the Discovery dataset. We next ran the platform on each of the five datasets independently (see **Methods**). Importantly, the platform identified zero models out of the 1822 with AUC-ROC and AUC-PR of above 0.8 across the five datasets, respectively, suggesting that the high performance of the 1822 models was not due to chance.

### Randomized data can be erroneously “classified” at reasonable AUC-ROC levels

The large number of models that resulted in a high AUC-ROC value were encouraging for the prospect of developing a blood-based classifier to supplement ASD diagnosis. However, depending on the model, a range of AUC-ROC values could be obtained. Thus, the question remained whether this range is significantly different from the AUC-ROC values that one could obtain from trying to classify subjects after randomizing their final diagnosis. To test this, we permuted the sample labels (i.e., ASD and TD) for all subjects in our Discovery set and ran the pipeline to test all feature engineering and classification methods. Importantly, we tested all 42,840 candidate models and found the median AUC-ROC score was 0.5101 with the 95th CI (0.4238-0.6498) on the randomized samples. Importantly, we found very rare but erroneous “classification” could be made with AUC-ROC values up to 0.8471 (see **Methods, Figure 1D**), which exceeded values reported for actual classifiers in the past. Thus, great care must be taken when developing classifiers for ASD to ensure the validity of a classifier.

### Classification is robust to sampling time, validation batches, and distinguishes ASD from language delay subjects

To assess the validity of the top-performing models and their reproducibility, we compared the performance of the 1822 models using two different datasets (i.e., Longitudinal, Replication datasets) that were not part of the discovery dataset nor used in any steps of feature route or classifier optimization(**Fig. 1C**), as further described below.

First, we evaluated the performance of the 1822 high-performing models on a Longitudinal dataset.The observed high reproducibility of model performance in Longitudinal samples suggests the existence of inherent biological signals in the ASD subjects that are independent of age or sample collection factors (see **Methods**, eResult 1 in Supplementary).

Second, we applied the 1822 models to an independent Replication dataset of 34 ASD and 31 TD. This dataset was assayed separately from the Discovery dataset and was independently analyzed. Notably, of 1822 models with AUC-ROC above 0.8 in the Discovery dataset, 1076 models (59.0%; Fisher’s Exact Test *P* < 2.2e-16) also had an AUC-ROC above 0.75 for the independent Replication ASD and TD samples (see **Methods**, eResult 2).

Third, we assessed the diagnostic specificity of the models by comparing performance of the 1822 models in separating a Diagnostic Specificity dataset of 24 LD toddlers from the Replication dataset. From 1822 models with AUC-ROC or AUC-PR above 0.8, none (Fisher’s Exact Test *P <* 2.2e-16) had an AUC-ROC or PR-ROC above 0.8 in this LD vs TD dataset. We also assessed the diagnostic specificity of the models by comparing performance of the 1822 models in separating a Diagnostic Specificity dataset of 24 LD toddlers from the ASD samples. From 1822 models with AUC-ROC or AUC-PR above 0.8, 26 (1.42%; Fisher’s Exact Test *P* < 2.2e-16) had an AUC-ROC above 0.75 in this LD vs TD dataset (see **Methods**, eResult 2 in Supplementary).

### Genes driving classification are largely immune-associated

We characterized the feature routes of 1076 models showing AUC-ROC above 0.8 in the Discovery dataset and above 0.75 in the independent Replication dataset. The 1076 models mostly originated from 191 feature routes. The mean variance of AUC-ROC scores by 12 classifiers on one route is 7.510e-04 (95% CI is [6.968e-04, 8.117e-04]).

To identify the biological processes that contribute to these 191 feature routes, we scored all genes in these routes based on the number of times they occurred in different 191 routes (Supplementary Table 2). By comparing the top five hundred genes selected by the routes, we found the genes selected by the feature routes had a larger impact on the model performance than the 12 classifiers. Different routes that largely overlapped in selected feature genes also showed highly correlated behavior across different classifier methods (see **Methods**, Supplementary Figures 5 and 6). We next calculated the enrichment of biological processes among top performing genes using g:Profiler. In this work, the 500 most common genes (Supplementary Table 2) were enriched in the KEGG pathways of inflammation/immune related disease, including Acute myeloid leukemia (KEGG: hsa05221), Measles infection (KEGG: hsa01524) and other significant pathways such as oxidative phosphorylation (KEGG: hsa04070), Ras signaling pathway (KEGG: has04014) and Wnt signaling pathway (KEGG: hsa04310). Similarly, enriched Gene Ontology terms for biological processes were associated with the down-regulation in blood inflammation/immune cell response, transcriptional gene regulation and response to cytokines, which were consistent with previous studies^30,34,36^ (see **Methods**, Supplementary Table 3).

**Table 2.** Distribution of Prenatal Events Among the Three Groups.

| Severe prenatal events | ASD<br>Above-the-mean | ASD Below-the-mean | TD |
| --- | --- | --- | --- |
| Total subjects (n) | 79 | 45 | 107 |
| Hospitalizations during pregnancy | 5 | 5 | 6 |
| Surgery during pregnancy | 1 | 3 | 4 |
| Confinement to bed during pregnancy | 8 | 8 | 9 |
| General Anesthesia during delivery | 10 | 11 | 10 |
| Total (%) | 19 (24.1) | 20 (44.4) | 24 (22.4) |
| **Negative control events** |  |  |  |
| Nausea | 4 | 2 | 12 |
| Morning sickness | 42 | 18 | 64 |
| Swelling | 23 | 10 | 26 |
| Total (%) | 52 (65.8) | 24 (53.3) | 70 (65.4) |

**ASD above-the-mean:** autism spectrum disorder with ensemble score over 0.714; **ASD below-the-mean:**
below 0.714; TD: typical development.

**Table 3.** Statistical Differences between the Three Groups.

|  | Odds ratio | *P* value |
| --- | --- | --- |
| **ASD above-the-mean vs. TD** | 0.914 | 0.861 |
| **ASD below-the-mean vs. TD** | 2.746 | 0.0102 |
| **ASD hard- vs. above-the-mean** | 2.506 | 0.027 |

795 *P* value is calculated by the two-sided Fisher's exact test

### High-performing models typically classify subjects similarly

We next leveraged our set of 1076 high-performing models to determine if there were different classification approaches that identify and separate ASD subjects into subgroups. To do so, we clustered the 175 subjects in the Discovery datasets based on classification score similarity. The classification scores provide a probabilistic measure on the certainty of a model on the diagnostic status of each discovery sample. It ranges from 0 to 1 with 0 being the highest certainty in TD status and 1 being the highest certainty in ASD status. The hierarchical clustering analysis found only one ASD group separated from TD subjects (**Figure 2A**). This shows that the 1076 models, overall, worked similarly in their assignment of classification scores to the subjects; that is, there was no evidence that different models selected different groups of ASD subjects. This was also true in analysis of the independent Replication dataset (**Figure 2A**).

**Figure 2.**
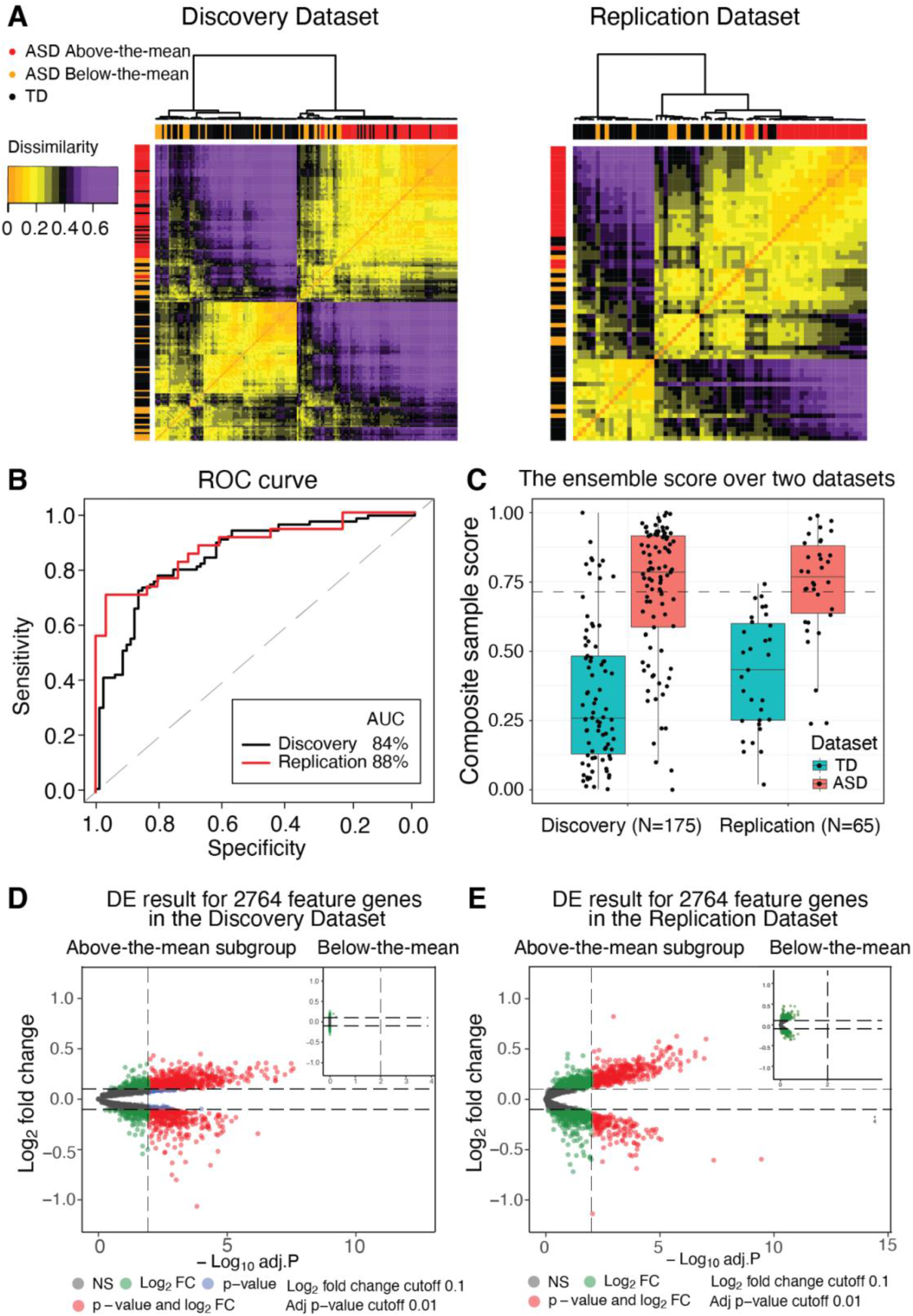
Blood transcriptome ASD subtypes identified by our classification platform. **A**. ASD and TD subjects show distinct classification patterns. The classification scores for each subject in the evaluation set is based on their score in the held-out sample. The red, orange and black bars on the sides represent above-the-mean ASD, below-the-mean ASD and TD subjects respectively. The orange and purple colors represent the gradient of dissimilarity between subjects based on their classification scores. **B**. The AUC-ROC results on the ensemble classification model generated by the Baysician model averaging approach. **C**. Subject ensemble scores based on the ensemble model. The quantile information of the main TD group is Min=0.0029, Q1=0.1337, Median=0.2568, Q3=0.4866, Max=0.9996; main ASD group is Min=0, Q1=0.576, Median=0.7859, Q3=0.9161, Max=1; Replication TD group is Min=0.0168, Q1=0.2454, Median=0.4196, Q3=0.5862, Max=0.7349; Replication ASD group is Min=0.238, Q1=0.628, Median=0.762, Q3=0.879, Max=0.990. **D**. and **E**. The differential expression analysis of 2764 protein coding feature genes. The volcano plots showed the adjusted *P* value (cutoff=0.01) vs logFold changes (cutoff=0.1) of genes in the above-the-mean to TD subjects and below-the-mean subjects to TD subjects in Discovery dataset and Replication dataset.

### Bayesian model averaging created a single ensemble classifier from the 1076 models

We next employed a Bayesian model averaging technique to assign an ensemble classification score to each subject. Each subject’s ensemble score was an average of his classification scores across the 1076 high-performing models weighted by the model performance as measured by AUC-ROC. We used this ensemble classifier model to calculate the AUC-ROC and AUC-PR values from the composite of the 1076 models. This analysis indicated an AUC-ROC of 84.42% and 87.85% for Discovery and Replication datasets, respectively (**Figure 2B**). We observed a similar performance based on PR curves with AUC-PR of 84.10% and 91.25% for the Discovery and Replication datasets, respectively, which is significantly higher than the baseline we created by using the naive Random Forest model with 72.32% AUC-ROC (ROC.test *P* < 0.01^44^) (see **Methods, eResult 3 in supplementary**).

Given that social attention abnormalities are well known in ASD, we enhanced the ensemble model by incorporating toddlers’ gaze fixation on non-social (i.e., geometric) images as measured in an eye-tracking test known as GeoPref test and used in previous studies^45–47^. The updated model was tested on 132 of 175 Discovery dataset subjects and 41of 65 Replication dataset subjects who had available Geo-Fixation data (e.g., moderate or good data quality, total looking time > 50%). By directly classifying the subjects who had percent fixation on non-social images >69% as ASD (GeoPref-subtype)^45–47^, the model AUC-ROC increased to 88.02% (AUC-PR 88.10%) and 89.42% (AUC-PR 92.11%) for Discovery and Replication datasets (see **Methods**, Supplementary Figure 7).

### ASD subjects with below-ASD-mean score were more likely to have had a prenatal risk event

Subsequently, we examined the clinical characteristics of ASD and TD subjects with different ensemble classifier scores. First, we stratified ASD toddlers by the mean ASD classifier score of 0.714 and grouped them into the above-the-mean ASD subgroup and the below-the-mean subgroup (**Figure 2C**). We next visualized the calculated ensemble score for each of 175 Discovery and 65 Replication datasets to better understand its power in distinguishing ASD from TD samples (**Figure 2C**).

The below-the-mean subgroup had no differential expressed (DE) genes compared with the TD group, while the above-the-mean ASD subgroup had 1162 DE genes. There were 394 DE genes in the first 500 gene features selected by the 191 feature routes and 700 DE genes in the first 1000 gene features demonstrating the ensemble model captured the dysregulation of gene expression pattern in the above-the-mean subgroup (Supplementary Table 2). After performing the DE gene analysis on the above-the-mean ASD subgroup vs TD, we conducted the enrichment analysis on the biological processes GO terms, and the pathway analysis with the KEGG database using g:Profiler^48,49^. In addition to the inflammation/immune response, transcriptional gene regulation, and response to cytokine, we found more gene ontology terms associated with the mitotic cell cycle. The significant pathways included the cell cycle (KEGG:hsa04110), PI3K-AKT (KEGG:hsa04151), RAS signaling pathways (KEGG:hsa04014), and Wnt signaling pathways (KEGG:hsa04310), which is consistent with our previous finding^42^ (Supplementary Table 3).

Diagnostic and psychometric scores were not significantly different between the above-the-mean ASD subgroup and the below-the-mean ASD subgroup (Supplementary Table 4). However, there were differences in prenatal risk factors, wherein ASD toddlers with classifier scores above the ASD mean had significantly fewer prenatal neurodevelopmental risk events, while ASD toddlers below the mean had disproportionately more prenatal risk scores than TD toddlers (**Table 2 and 3**). We tested if there was a different ratio of severe prenatal events that could potentially impact ASD development between the two transcriptome ASD subgroups. We found a similar rate of prenatal events between TD subjects and above-the-mean ASD subjects (Odds Ratio: 0.88, Fisher’s Exact Test *P* = 0.84). However, there was a significant enrichment of prenatal events among the below-the-mean ASD subjects compared to TD subjects (Odds Ratio: 2.78; Fisher’s Exact Test *P* = 0.013). As a negative control, prenatal events that are unlikely to affect ASD development were not enriched among ASD subjects with below the ASD mean score. These results suggest the possible existence of different underlying etiological factors between ASD subjects with above vs below the mean ASD classifier score.

Overall, while 83 (64.5%) of ASD subjects showed an ensemble classification score of above 0.714, only 11 (9.73%) of TD subjects reached this score, underscoring the high performance of the ensemble score in separating ASD and TD cases. This indicates higher ensemble diagnostic scores are strongly driven by significantly DE genes in leukocytes that enrich the GO biological processes described above and in Supplementary table 3, while DE genes are less of a classifier factor in ASD subjects with lower classification ensemble scores.

## Discussion

Despite its prenatal beginning^1–16^, ASD diagnosis remains behavior-based and the median age of first diagnosis is about 52 months. Partially due to its genetic and clinical heterogeneity, no single diagnostic marker has been found that can accurately and reproducibly diagnose more than a small subset of affected children. Even among those capable of correctly diagnosing small subsets of ASD infants and toddlers, few have proven clinically useful, cost-effect, and/or practical at the ages when early detection and diagnosis are most needed and could be mostly important for the child and family^47^.

We used a unique approach to this dilemma: We addressed ASD genetic and clinical heterogeneity with classifier heterogeneity. That is, since we expected heterogeneity in classifier gene features, we designed a classifier pipeline to train, test, and validate 42,840 models generated from 3,570 gene feature routes and 12 classification methods to correctly identify ASD at ages 1 to 4 years. Then, rather than selecting a single “best” performing model, we used Bayesian model averaging to bring together 1,076 “heterogeneous” high-performing models involving 191 different feature routes and 2,764 gene features.

This approach produced a composite “ensemble” model: (1) having one of the highest performances (84%-88%) yet reported for large samples of both Discovery and Replication subjects; (2) having stability across subjects; (3) performing well in both ASD vs TD toddlers and ASD vs LD toddlers; and (4) performing accurately across the ASD spectrum from more affected to less affected.

Moreover, this composite ensemble model incorporates both differentially expressed (DE) genes and non-DE genes. This may be relevant to the known complexity of ASD genetics, which may involve common and rare variants and any one or more of >200 different ASD risk gene mutations in different individuals. Non-genetic heterogeneity was also detected here insofar as those with ASD classifier scores below the overall ASD mean tended to have more prenatal risk events in their history than those ASD toddlers with above the mean scores. This opens the important potential to utilize these ASD ensemble classifier scores in future research to home in ASD subtypes more likely to be principally driven by genetic versus subtypes more likely to be driven by non-genetic or a combination of non-genetic and genetic factors.

Our ensemble features include genes with PI3K-AKT, RAS, and Wnt signaling pathways, immune/inflammation, response to cytokines, transcriptional regulation, and mitotic cell cycle. Meta-analysis, review, and original data papers on ASD blood gene expression^29–39^ generally show that common ground across many studies is dysregulation in PI3K-AKT-mTOR and RAS signaling pathways, immune/inflammation processes, the KEGG natural killer cell pathway, cell cycle, apoptosis, neurogenesis as well as increased ribosomal translation signal. These main signaling pathways, immune and cell cycle correspondences are notable despite the fact that (1) across more than 35 studies diverse blood-based samples were examined (whole blood, lymphoblastoid cell lines, peripheral blood mononuclear cells, or other types of leukocyte capture methods); (2) most studies did not actively account for sex-related, age-related or clinical-symptom heterogeneity as moderating factors; (3) 84% of them had less than 100 ASD subjects averaging 28 subjects/study; and (4) most studies focused on older ASD children and adults ^29–39^.

PI3K-AKT, RAS and Wnt signaling pathways may be pivotal to ASD prenatal neural maldevelopment. Recently, in a large sample study, we discovered that ASD toddlers had significant upregulation of PI3K-AKT, RAS-ERK and Wnt signaling pathways in both leukocytes and iPSC-derived prenatal neural progenitors and neurons^42^. This leukocyte dysregulation in ASD toddlers aged 1-4 years olds correlated with ASD social symptom severity^42^. Moreover, these pathways in leukocytes are downstream targets of regulatory risk ASD genes^42^.

Leukocyte gene expression has high potential not only for objective molecular diagnostic identification of ASD at 1 and 2-year-olds of age, but also for understanding ASD genetic and clinical heterogeneity at early ages. For example, leukocyte expression signatures correlate with brain size in ASD^29^, and to atypical cortical patterning subtypes ASD toddlers with poor language outcome^50^. Leukocyte expression also relates to hypoactivation response to affective speech in ASD toddlers with poor language outcome^51^. Finally, we found multivariate leukocyte expression signatures can predict trajectories of response to early intervention treatment^52^, which underscores the mechanistic relevance of leukocytes to ASD and clinically important phenomena that can be individualized to specific patients.

In sum, a large amount of literature, meta-analyses, and the robust leukocyte diagnostic discoveries in the present study, all point to the importance of leukocyte cell biology in ASD and show that ASD-relevant dysregulated gene expression is not restricted to the brain. Indeed, the earliest prenatal drivers of deviant ASD development are broadly expressed regulatory genes, and such genes are active not only in the brain but also in leukocytes and other organs^1–3,29,30,42,51,53^. Over 60% of ASD risk genes are broadly expressed regulatory genes^1,3,16^, and 97 of the top broadly expressed regulatory ASD risk genes express in leukocytes as well as the brain. Thus, broadly expressed genes that constitute the majority of ASD risk genes are upregulated in the early prenatal life, which impact multiple stages of prenatal brain development from 1^st^ and 2^nd^ trimester proliferation and neurogenesis through neurite outgrowth and synaptogenesis in the 3^rd^ trimester^1,3,16^. They do so by disrupting gene expression in signaling pathways, such as ASD-important PI3K-AKT, RAS-ERK and Wnt signaling pathways^1,3,42^, which further disrupt prenatal functions. ASD leukocyte expression has high relevance to ASD diagnostic, structural, and neural functional phenotypes and clinically different prognostic endpoints^50,51^.

Here we developed an innovative high-performing ASD molecular classifier with heterogeneous gene features designed to address ASD genetic and clinical heterogeneity. This high-level performance in ASD male toddlers aged 1 to 4-year-olds opens the possibility of further refining ASD molecular classifiers optimized for ethnicity, age, and sex. The ensemble gene expression ASD classifier reported here is enriched in gene features known to be involved in ASD prenatal and postnatal pathobiology, and as such, it succeeds because of this. Thus, it is more than a signal capable of classifying ASD. Instead, it is a signal of the underlying pathobiological bases of the disorder in a majority of affected toddlers and therefore has implications for future research targeting treatment-relevant mechanisms.

## Data Availability

All data have been archived at the NIMH NDA

## Acknowledgements

This work was supported by NIMH grant no. R01-MH110558 (E.C., N.E.L.), NIMH grant no. R01-MH080134 (K.P.), NIMH grant no. R01-MH104446 (K.P.), an NFAR grant (K.P.), NIMH grant no. P50-MH081755 (E.C.), and generous funding from the Novo Nordisk Foundation through the Center for Biosustainability at the Technical University of Denmark (grant no. NNF10CC1016517 to N.E.L.).

## Author Contributions

V.H.G., B.B., R.K., A.W.T.C, K.P., K.R., E.C. and N.E.L. conceived the project and designed the experiments. Y.X. and S.N. collected the samples, conducted transcriptome assays and managed the data. V.H.G., B.B. and R.K. analyzed the data. V.H.G., B.B., Y.X., R.K., K.P., E.C. and N.E.L. interpreted the results and wrote the manuscript. E.C. and N.E.L. supervised the project.

## Competing interests

The authors declare no competing interests.

## Code Availability

We provide the code library in R and Python described in this work through Github: https://github.com/LewisLabUCSD/autism_classifier. We provide jupyter notebooks in python and R to generate our figures and analysis.

## METHODS

### Participant recruitment and clinical evaluation

Participants in this study included 264 male toddlers ages 1 to 4 years (**Table 1**). About 70% of toddlers were recruited from the general population as young as 12 months using an early screening, detection, and diagnosis strategy called the Get SET Early procedure^22^. Using this approach, toddlers who failed a broadband screen, i.e., the CSBS IT Checklist^23^, at well-baby visits in the general pediatric community settings were referred to our center for a comprehensive evaluation. The remaining subjects were obtained by general community referrals. All toddlers received a battery of standardized psychometric tests by highly experienced Ph.D.-level psychologists, including the Autism Diagnostic Observation Schedule (ADOS; Module T, 1 or 2), the Mullen Scales of Early Learning, and the Vineland Adaptive Behavior Scales. Testing sessions routinely lasted 4 hours in one day or occurred across 2 separate days. Toddlers younger than 36 months upon initial clinical evaluation were followed longitudinally approximately every 9 months until final diagnosis at ages 2-4 years. Most toddlers were tracked and diagnosed using the appropriate module of the ADOS (i.e., ADOS Module-Toddler, Module-1, or Module-2) between 24-49 months, an age when the diagnosis of ASD is relatively stable^24^; 264 toddlers had their final diagnostic evaluation between 18 to 24 months. 127 toddlers were ASD, 113 TD, 24 LD (**Table 1**). The 264 toddlers included in the current study were among those in a 1,269 subjects study of early diagnostic stability in ASD^43^. Research procedures were approved by the Institutional Review Board of the University of California, San Diego. Parents of subjects underwent Informed Consent Procedures with a psychologist or study coordinator at the time of their child’s enrollment.

### Blood sample collection

Transcriptome data based on microarrays included previously published samples^25^ and 46 novel samples. For these, blood samples were collected from each of the 264 subjects; longitudinal blood samples were collected from a subset of 33 subjects (**Table 1**). They were usually taken at the end of clinical evaluation sessions. To monitor health status, the temperature of each toddler was monitored using an ear digital thermometer immediately preceding the blood draw. The blood draw was scheduled for a different day when the temperature was higher than 99 Fahrenheit. Moreover, the blood draw was not taken if a toddler had some illness (e.g., cold or flu), as observed by us or stated by parents. We collected four to six milliliters of blood into ethylenediaminetetraacetic-coated tubes from all toddlers. Blood leukocytes were captured and stabilized by LeukoLOCK filters (Ambion) and were immediately placed in a −20°C freezer. Total RNA was extracted following standard procedures and manufacturer’s instructions (Ambion).

### Data processing and differential gene expression analysis of the microarray primary dataset

From each blood sample, leukocyte RNA was extracted, and gene expression was assayed using Illumina HT-12 platform. All arrays were scanned with the Illumina BeadArray Reader and read into Illumina GenomeStudio software (version 1.1.1). Raw Illumina probe intensities were converted to expression values using the lumi package^54^. We employed a three-step procedure to filter for probes with reliable expression levels. First, we only retained probes that met the detection *P* value<0.05 cut-off threshold in at least 3 samples. Second, we required probes to have expression levels above the 95^th^ percentile of negative probes in at least 50% of samples. The probes with detection *P* value>0.1 across all samples were selected as negative probes and their expression levels were pooled together to estimate the 95^th^ percentile expression level. Third, for genes represented by multiple probes, we considered the probe with the highest mean expression level across our dataset, after quantile normalization of the data. These criteria led to the selection of 14,312 coding genes as expressed in our leukocyte transcriptome data, which highly overlaps with the reported estimate of 14,555 protein-coding genes (chosen based on unique Entrez gene IDs) for whole blood by the GTEx consortium^55^.

### Building the classifier platform on the Discovery set

The Discovery dataset included 175 subjects with 14,132 gene features. The pipeline ran five iterations. At the beginning of each iteration, the pipeline held out 20% of samples and used the remaining 80% of samples for hyper-parameter selection, feature selection, and classifier training. In the first step, feature filtration, five methods were used (Supplementary Figure 1), including no (no action), cov (remove 50% feature with the smaller coefficient of variation), var (remove 50% feature with smaller variance), cov_var (remove 50% feature with the smaller coefficient of variation and then remove 50% feature with smaller variance in the rest), varImportance (keep only the 25% features with the highest variance).

The second step, feature selection, includes seven groups of methods (Supplementary Figure 1) with 102 methods in total. The methods within each group are conceptually similar, but use different approaches. These groups are no (no action), grn^56^ (genetic regulatory network), z-score, selectV^57^, svm^58^, GSEA^59^, DE-analysis^60^. The Grn group has 14 methods, based on gene regulatory network analysis. The Z-score group has 4 methods for selecting features with high z-score variance. It picks the features with variance in the first quartile and then selects features within the 3rd quartile of the context likelihood of relatedness. The SelectV group has 54 methods and bases on the “Variable Selection for High-Dimensional Supervised Classification”^57^. In this group, features are first filtered by ‘ExpHC’,’HC’, ‘Fair’ methods (with/without comvar). The remaining features are further selected by the grn method with different parameters. The Svm group has 4 methods leveraging support vector machines with feature selection methods “scad”, “L1”, “ElasticNet” and “scad+L2”. The GSEA group has 10 methods based on gene set enrichment analysis. The features are chosen with either FDR (False discovery rate)<0.1 or FDR<0.25. The features are ordered by FDR value within both feature groups and selected by choosing the lowest 10%, lowest 20%, lowest 50%, or below mean threshold. The remaining features are filtered by the cor and the grn3 method. The DE-analysis group has 15 methods and is picking features based on differential gene expression. The methods pick features with the combination of *P* value (*P* < 0.05 or *P* <0.01), fold change (logFc >0 or logFc<0 or both), and different numbers of features (1000, 500, or 100 features with the lowest *P*).

The third step was feature reduction (Supplementary Figure 1). Seven methods were used: no (no feature reduction), WGCNA^61^, logisticFwd, SIS^62^, principal component regression (PCR)^63^, partial least squares regression (PLSR)^64^, canonical powered partial least squares (CPPLS)^64^. The WGCNA, weighted correlation network analysis, calculates modules of co-expressed genes^61^, and we select hub genes from each module. The LogisticFwd method deploys stepwise logistic regression with the forward method to select features. The SIS^62^ method returns the features selected by the SIS R package, which implements the “Iterative Sure Independence Screening” for selecting variants. PCR regresses on a subset of principal components for dimensionality reduction, while PLSR finds a dimensionality-reducing linear regression model by projecting the variables to a new space, and CPPLS allows for discrete and continuous responses in the PLS model. After three steps, up to 1320 gene routes were created that can be used in the classification step.

The classification step exploited 12 classifiers (Supplementary Figure 1), including reg (linear model), logReg^65^ (logistic regression), lda^65^ (Linear Discriminant Analysis), qda^65^ (Quadratic Discriminant Analysis), ridgeReg^66^ (GLM with ridge regularization), lassoReg^66^ (GLM with lasso regularization), ridgeLogReg^66^ (logistic regression with ridge regularization), lassoLogReg^66^ (logistic regression with lasso regularization), elasticNetLogReg^66^ (logistic regression with elastic net regularization), boosting^67^ (Generalized Boosted Regression Modeling with Bernoulli distribution), randomForest^68^ (random forest) and bagging^68^ (random forests with bagging to reduce the complexity). After training a classifier, the diagnostic ability was evaluated by AUC-PR (precision-recall) curve and AUC-ROC (Receiver operating characteristic) curve^44,69^.

After five iterations, the averaged value of AUC area under precision=0.95 and precision=0.85 was used to compare the feature route and classifier performance. 1822 (11.5%) models with classification AUC-ROC of above 0.8 were found.

### Label permutation data

To generate the randomized background, we shuffled the diagnostic label of the Discovery dataset and randomly separated the data into training/validation segments (85%/15%). Then we perform the 5-fold cross validation on the permuted dataset.

### Random forest model created the baseline AUC-ROC

Used 175 Discovery and 65 Replication dataset. Data are prepared by randomly splitting the Independent Replication dataset into a separate validation and test set. The same Discovery dataset is used for training. Using the Discovery set, we perform feature selection for the top ***q*** variables, as scored by the chi-squared statistic. From a grid, ***q*** is chosen based on which grid value yields the highest mean AUC after 100 random forest evaluations on the validation set. Additionally, we average the feature importance rankings over the 100 iterations to provide a more comprehensive view of important genes. With the selected ***q*** value, we train a random forest classifier and evaluate on the held-out test set.

### Visualization of the classifier AUC-ROC scores vs route mean AUC-ROC score

The 12-classifier mean AUC-ROC scores of 1320 routes are calculated and set as x-axis. 12 classifiers’AUC-ROC scores are visualized as y-axis (Supplementary Figure 3C and D). Then, the 12-classifiers mean AUC-ROC scores of 1320 routes are binned into 10 quantiles and the variance of the 12-classifiers AUC-ROC score for each route are calculated and presented as boxplot (Supplementary Figure 3E and F).

### Reproducibility on Longitudinal dataset

Models were trained on the Discovery dataset (175 subjects) and then tested on the Longitudinal dataset (Supplementary Figure 2). The Longitudinal dataset contained 18 ASD and 15 TD toddlers in the Discovery set. 1656 (90.8%) models showed AUC-ROC of >0.8 in the Longitudinal dataset.

### Validation of Independent Replication dataset

The independent Replication dataset consisted of 89 subjects (34 ASD, 31 TD, and 24 LD). To test the sensitivity, the models were trained on the Discovery dataset (175 subjects together) and then tested on the dataset with 34 ASD and 31 TD. The AUC-ROC values were used to evaluate the performance of the selected 1822 high-performing models. 1076 models (59.0%) had an AUC-ROC value greater than 0.75 in the Replication dataset

### Bayesian model averaging to create a single transcriptomic ensemble classifier from the 1822 models

After training on the Discovery dataset, the 1076 models that also had AUC-ROC value >0.8 on the Replication dataset were selected. The ensemble score was the sum of weighted predictions of selected models. The weight is the mathematical average of the square of (AUC-ROC value minus 0.7). In a model selection, we use training data D to select a good model M (according to some score) to use in predicting a targeted outcome T of interest based on patient features X, namely, P(T | X, M). BMA is based on the notion of averaging over a set of possible models and weighting the prediction of each model according to its probability given training data D, as shown in equations.

- 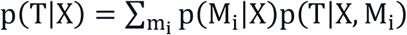 M is the model, T is the prediction and X is the data.
- 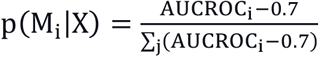

The ensemble scores of the independent dataset are calculated based on the same model built. The scores are then rescaled to 0 and 1.

- 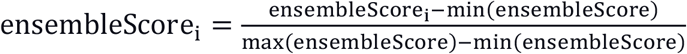

### The gene set similarity score between routes

We compared the first five hundred genes that existed in the 191 routes (Supplementary Table 2). In order to retrieve the genes from dimension deduction methods such pcr, cppls and plsr, we calculated the feature weights and the gene weights in each feature. Then, we ranked the genes with the weights they contribute to all features and selected the genes that cumulated to 66% of total weights.

- 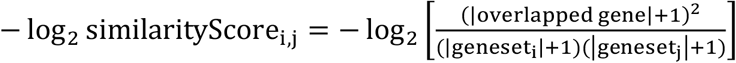

In the Supplementary Figure 5 and 6, the distance of the routes gene similarity is presented as −log_2_ similarityScore_i,j_.

### Calculating the biological enrichment of DE genes between above-mean ASD and TD

After generating the prediction score by the ensemble classifier, we stratified ASD toddlers in the Discovery dataset into those with above and below the Discovery dataset’s ensemble classifier mean score of 0.714. Those above this threshold were considered “above-the-mean” and below 0.714 are considered “below-the-mean”. Then we conducted differential expression (DE) analysis on the above-the-mean group (n=59) subjects and TD (n=82) subjects. Then we conducted differential expression (DE) analysis on the below-the-mean group (n=34) and TD (n=82) subjects. The limma package^60,70^ was then applied on quantile-normalized data for differential expression analysis in which moderated t-statistics were calculated by robust empirical Bayes methods. We use adj.p.value<0.01 and logFC>0.1 to select genes and generate the volcano plot. The Gene Ontology (GO) enrichment was conducted using g:Profiler^48^ (https://biit.cs.ut.ee/gprofiler/gost) with 12695 protein-coding genes (12695/14132 gene features) as background (g:Profiler, advanced option/statistical domain scope: Custom; custom over annotated genes). We only checked the “GO biological process” and KEGG terms of size 15-1500 in the biological process. The threshold is “Significance threshold: B-H FDR<0.1”. Then the terms are clustered with REVIGO^71^, ordered with *P* value (http://revigo.irb.hr/). The connections across terms are visualized by the Cytoscape 3.8.2^72^.

### Clinical and prenatal characteristics associated with ASD classifier scores

To examine clinical and prenatal characteristics associated with ensemble classifier scores, we stratified 127 ASD toddlers in the Discovery and independent Replication datasets into those with above and below the score of 0.714 (again as the easy and the hard group). The t-test is used to compare the diagnostic and psychometric scores between ASD toddlers among two groups (**Table 3**). Among 127 subjects, 124 subjects have complete prenatal records. The t-test was used to compare the diagnostic and psychometric scores between ASD toddlers among two groups (**Table 3**). Among 127 subjects, 124 subjects had complete prenatal records. The fisher.t.test was used to compare the prenatal risk factors across easy, hard and TD groups. Next, we stratified ASD toddlers based on the group means of CoSo Total symptom severity and Mullen cognitive scores. As shown in Supplementary Table 2, these clinical stratifiers also did not differentiate ASD toddlers with higher or lower ensemble classifier scores. Thus, the ensemble classifier was equally effective as a diagnostic tool across a considerable range of early-age ASD diagnostic symptoms and cognitive presentation.

We performed analogous stratifications within the TD Discovery group and found no ADOS or Mullen differences between higher or lower than the TD mean ensemble classifier score, nor differences in the ensemble scores of TD toddlers with high vs lower diagnostic and psychometric scores (Supplementary Table 2).

### Geo Fixation (non-social) characteristics associated with ASD classifier scores

After we have the ensemble score, we tested the ensemble model on 132/175 of the subjects found in the Discovery dataset that have high quality Geo Fixation (non-social) data and 41/65 of the subjects found in the independent Replication dataset. The rule is

- 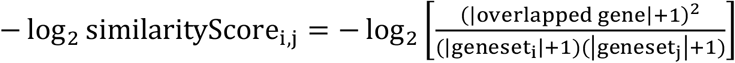
- ensembleScore_i_= 1 if geo-fixation percentage>69%, else ensembleScore_i_

## Notes

### Competing Interest Statement

The authors have declared no competing interest.

### Author Declarations

UC San Diego IRB Project #202115X Discovering Biomarkers, Causes and Treatment of ASD through Clinical and Biological Studies

## Reference

1. Courchesne E, Gazestani VH, Lewis NE. Prenatal origins of ASD: The when, what, and how of ASD development. Trends Neurosci. 2020;43(5):326–342.

2. Courchesne E, Pramparo T, Gazestani VH, Lombardo MV, Pierce K, Lewis NE. The ASD Living Biology: from cell proliferation to clinical phenotype. Mol Psychiatry. 2019;24(1):88–107.

3. Gazestani V, Chiang AWT, Courchesne E, Lewis NE. Autism genetics perturb prenatal neurodevelopment through a hierarchy of broadly-expressed and brain-specific genes. bioRxiv. Published online 2020.

4. Courchesne E, Mouton PR, Calhoun ME, et al. Neuron number and size in prefrontal cortex of children with autism. JAMA. 2011;306(18):2001–2010.

5. Marchetto MC, Belinson H, Tian Y, et al. Altered proliferation and networks in neural cells derived from idiopathic autistic individuals. Mol Psychiatry. 2017;22(6):820–835.

6. Courchesne E, Pierce K. Why the frontal cortex in autism might be talking only to itself: local over- connectivity but long-distance disconnection. Curr Opin Neurobiol. 2005;15(2):225–230.

7. Willsey AJ, Sanders SJ, Li M, et al. Coexpression networks implicate human midfetal deep cortical projection neurons in the pathogenesis of autism. Cell. 2013;155(5):997–1007.

8. Courchesne E, Pierce K, Schumann CM, et al. Mapping early brain development in autism. Neuron. 2007;56(2):399–413.

9. Stoner R, Chow ML, Boyle MP, et al. Patches of Disorganization in the Neocortex of Children with Autism. New England Journal of Medicine. 2014;370(13):1209–1219. doi:10.1056/nejmoa1307491

10. Parikshak NN, Luo R, Zhang A, et al. Integrative Functional Genomic Analyses Implicate Specific Molecular Pathways and Circuits in Autism. Cell. 2013;155(5):1008–1021. doi:10.1016/j.cell.2013.10.031

11. Packer A. Neocortical neurogenesis and the etiology of autism spectrum disorder. Neurosci Biobehav Rev. 2016;64:185–195.

12. Kaushik G, Zarbalis KS. Prenatal Neurogenesis in Autism Spectrum Disorders. Frontiers in Chemistry. 2016;4. doi:10.3389/fchem.2016.00012

13. Krishnan A, Zhang R, Yao V, et al. Genome-wide prediction and functional characterization of the genetic basis of autism spectrum disorder. Nat Neurosci. 2016;19(11):1454–1462.

14. Donovan APA, Basson MA. The neuroanatomy of autism - a developmental perspective. J Anat. 2017;230(1):4–15.

15. Grove J, Ripke S, Als TD, et al. Identification of common genetic risk variants for autism spectrum disorder. Nat Genet. 2019;51(3):431–444.

16. Satterstrom FK, Kosmicki JA, Wang J, et al. Large-Scale Exome Sequencing Study Implicates Both Developmental and Functional Changes in the Neurobiology of Autism. Cell. 2020;180(3):568-584.e23.

17. Bai D, Yip BHK, Windham GC, et al. Association of Genetic and Environmental Factors With Autism in a 5-Country Cohort. JAMA Psychiatry. 2019;76(10):1035–1043.

18. Bal VH, Kim S-H, Fok M, Lord C. Autism spectrum disorder symptoms from ages 2 to 19 years: Implications for diagnosing adolescents and young adults. Autism Res. 2019;12(1):89–99.

19. Bacon EC, Courchesne E, Barnes CC, et al. Rethinking the idea of late autism spectrum disorder onset. Dev Psychopathol. 2018;30(2):553–569.

20. Bacon EC, Osuna S, Courchesne E, Pierce K. Naturalistic language sampling to characterize the language abilities of 3-year-olds with autism spectrum disorder. Autism. 2019;23(3):699–712.

21. Autism and Developmental Disabilities Monitoring Network Surveillance Year 2006 Principal Investigators, Centers for Disease Control and Prevention (CDC). Prevalence of autism spectrum disorders - Autism and Developmental Disabilities Monitoring Network, United States, 2006. MMWR Surveill Summ. 2009;58(10):1–20.

22. Baio J, Wiggins L, Christensen DL, et al. Prevalence of Autism Spectrum Disorder Among Children Aged 8 Years - Autism and Developmental Disabilities Monitoring Network, 11 Sites, United States, 2014. MMWR Surveill Summ. 2018;67(6):1–23.

23. Christensen DL, Braun KVN, Baio J, et al. Prevalence and Characteristics of Autism Spectrum Disorder Among Children Aged 8 Years - Autism and Developmental Disabilities Monitoring Network, 11 Sites, United States, 2012. MMWR Surveill Summ. 2018;65(13):1–23.

24. Maenner MJ, Shaw KA, Baio J, et al. Prevalence of Autism Spectrum Disorder Among Children Aged 8 Years - Autism and Developmental Disabilities Monitoring Network, 11 Sites, United States, 2016. MMWR Surveill Summ. 2020;69(4):1–12.

25. Lombardo MV, Lai M-C, Baron-Cohen S. Big data approaches to decomposing heterogeneity across the autism spectrum. Mol Psychiatry. 2019;24(10):1435–1450.

26. Robinson EB, St Pourcain B, Anttila V, et al. Genetic risk for autism spectrum disorders and neuropsychiatric variation in the general population. Nat Genet. 2016;48(5):552–555.

27. Ahn K, An SS, Shugart YY, Rapoport JL. Common polygenic variation and risk for childhood-onset schizophrenia. Mol Psychiatry. 2016;21(1):94–96.

28. Clarke T-K, Lupton MK, Fernandez-Pujals AM, et al. Common polygenic risk for autism spectrum disorder (ASD) is associated with cognitive ability in the general population. Mol Psychiatry. 2016;21(3):419–425.

29. Pramparo T, Lombardo MV, Campbell K, et al. Cell cycle networks link gene expression dysregulation, mutation, and brain maldevelopment in autistic toddlers. Mol Syst Biol. 2015;11(12):841.

30. Pramparo T, Pierce K, Lombardo MV, et al. Prediction of autism by translation and immune/inflammation coexpressed genes in toddlers from pediatric community practices. JAMA Psychiatry. 2015;72(4):386–394.

31. Ch’ng C, Kwok W, Rogic S, Pavlidis P. Meta-Analysis of Gene Expression in Autism Spectrum Disorder. Autism Res. 2015;8(5):593–608.

32. Diaz-Beltran L, Esteban FJ, Wall DP. A common molecular signature in ASD gene expression: following Root 66 to autism. Transl Psychiatry. 2016;6:e705.

33. Tylee DS, Hess JL, Quinn TP, et al. Blood transcriptomic comparison of individuals with and without autism spectrum disorder: A combined-samples mega-analysis. Am J Med Genet B Neuropsychiatr Genet. 2017;174(3):181–201.

34. He Y, Zhou Y, Ma W, Wang J. An integrated transcriptomic analysis of autism spectrum disorder. Sci Rep. 2019;9(1):11818.

35. Lee SC, Quinn TP, Lai J, et al. Solving for X: Evidence for sex-specific autism biomarkers across multiple transcriptomic studies. Am J Med Genet B Neuropsychiatr Genet. 2019;180(6):377–389.

36. Kong SW, Collins CD, Shimizu-Motohashi Y, et al. Characteristics and predictive value of blood transcriptome signature in males with autism spectrum disorders. PLoS One. 2012;7(12):e49475.

37. Gregg JP, Lit L, Baron CA, et al. Gene expression changes in children with autism. Genomics. 2008;91(1):22–29.

38. Enstrom AM, Lit L, Onore CE, et al. Altered gene expression and function of peripheral blood natural killer cells in children with autism. Brain Behav Immun. 2009;23(1):124–133.

39. Ansel A, Rosenzweig JP, Zisman PD, Melamed M, Gesundheit B. Variation in Gene Expression in Autism Spectrum Disorders: An Extensive Review of Transcriptomic Studies. Front Neurosci. 2016;10:601.

40. Hyde KK, Novack MN, LaHaye N, et al. Applications of Supervised Machine Learning in Autism Spectrum Disorder Research: a Review. Review Journal of Autism and Developmental Disorders. 2019;6(2):128–146.

41. Hewitson L, Mathews JA, Devlin M, Schutte C, Lee J, German DC. Blood biomarker discovery for autism spectrum disorder: A proteomic analysis. PLoS One. 2021;16(2):e0246581.

42. Gazestani VH, Pramparo T, Nalabolu S, et al. A perturbed gene network containing PI3K-AKT, RAS-ERK and WNT-β-catenin pathways in leukocytes is linked to ASD genetics and symptom severity. Nat Neurosci. 2019;22(10):1624–1634.

43. Pierce K, Gazestani VH, Bacon E, et al. Evaluation of the Diagnostic Stability of the Early Autism Spectrum Disorder Phenotype in the General Population Starting at 12 Months. JAMA Pediatr. 2019;173(6):578–587.

44. Robin X, Turck N, Hainard A, et al. pROC: an open-source package for R and S+ to analyze and compare ROC curves. BMC Bioinformatics. 2011;12:77.

45. Pierce K, Conant D, Hazin R, Stoner R, Desmond J. Preference for geometric patterns early in life as a risk factor for autism. Arch Gen Psychiatry. 2011;68(1):101–109.

46. Moore A, Wozniak M, Yousef A, et al. The geometric preference subtype in ASD: identifying a consistent, early-emerging phenomenon through eye tracking. Mol Autism. 2018;9:19.

47. Pierce K, Marinero S, Hazin R, McKenna B, Barnes CC, Malige A. Eye Tracking Reveals Abnormal Visual Preference for Geometric Images as an Early Biomarker of an Autism Spectrum Disorder Subtype Associated With Increased Symptom Severity. Biol Psychiatry. 2016;79(8):657–666.

48. Raudvere U, Kolberg L, Kuzmin I, et al. g:Profiler: a web server for functional enrichment analysis and conversions of gene lists (2019 update). Nucleic Acids Res. 2019;47(W1):W191-W198.

49. Kanehisa M, Goto S. KEGG: kyoto encyclopedia of genes and genomes. Nucleic Acids Res. 2000;28(1):27–30.

50. Lombardo MV, Eyler L, Pramparo T, et al. Atypical genomic patterning of the cerebral cortex in autism with poor early language outcome. doi:10.1101/2020.08.18.253443

51. Lombardo MV, Pramparo T, Gazestani V, et al. Large-scale associations between the leukocyte transcriptome and BOLD responses to speech differ in autism early language outcome subtypes. Nat Neurosci. 2018;21(12):1680–1688.

52. Lombardo MV, Busuoli EM, Schreibman L, et al. Pre-treatment clinical behavioral and blood leukocyte gene expression patterns predict rate of change in response to early intervention in autism. medRxiv. Published online 2020. https://www.medrxiv.org/content/10.1101/2020.12.21.20248674v1.abstract

53. Lombardo MV, Eyler L, Pramparo T, Gazestani VH. Atypical genomic cortical patterning in autism with poor early language outcome. bioRxiv. Published online 2021. https://www.biorxiv.org/content/10.1101/2020.08.18.253443v3.abstract

54. Du P, Kibbe WA, Lin SM. lumi: a pipeline for processing Illumina microarray. Bioinformatics. 2008;24(13):1547–1548. doi:10.1093/bioinformatics/btn224

55. Ardlie KG, Deluca DS, Segre AV, et al. The Genotype-Tissue Expression (GTEx) pilot analysis: Multitissue gene regulation in humans. Science. 2015;348(6235):648–660.

56. Meyer PE, Lafitte F, Bontempi G. minet: A R/Bioconductor package for inferring large transcriptional networks using mutual information. BMC Bioinformatics. 2008;9:461.

57. Antonio Pedro Duarte Silva. SelectV: Variable selection for high-dimensional supervised… In HiDimDA: High dimensional Discriminant Analysis. Published October 19, 2015. Accessed May 21, 2021. https://rdrr.io/cran/HiDimDA/man/SelectV.html

58. penalizedSVM: Feature Selection SVM using Penalty Functions. Accessed June 29, 2021. https://cran.r-project.org/web/packages/penalizedSVM/index.html

59. Subramanian A, Tamayo P, Mootha VK, et al. Gene set enrichment analysis: a knowledge-based approach for interpreting genome-wide expression profiles. Proc Natl Acad Sci U S A. 2005;102(43):15545–15550.

60. Ritchie ME, Phipson B, Wu D, et al. limma powers differential expression analyses for RNA-sequencing and microarray studies. Nucleic Acids Research. 2015;43(7):e47–e47. doi:10.1093/nar/gkv007

61. Langfelder P, Horvath S. WGCNA: an R package for weighted correlation network analysis. BMC Bioinformatics. 2008;9:559.

62. Saldana DF, Feng Y. SIS: An R Package for Sure Independence Screening in Ultrahigh-Dimensional Statistical Models. Journal of Statistical Software, Articles. 2018;83(2):1–25.

63. Mevik B-H, Wehrens R. Introduction to the pls Package. Help Section of The “Pls” Package of R Studio Software; R Foundation for Statistical Computing: Vienna, Austria. Published online 2015:1-23.

64. Wehrens R, Mevik B-H. The pls package: principal component and partial least squares regression in R. Published online 2007. https://repository.ubn.ru.nl/bitstream/handle/2066/36604/36604.pdf

65. Ripley BD. Modern Applied Statistics with S. springer; 2002.

66. Friedman J, Hastie T, Tibshirani R. Regularization paths for generalized linear models via coordinate descent. J Stat Softw. 2010;33(1):1–22.

67. Ridgeway G. Generalized Boosted Models: A guide to the gbm package. Update. 2007;1(1):2007.

68. Liaw A, Wiener M, Others. Classification and regression by randomForest. R news. 2002;2(3):18–22.

69. Grau J, Grosse I, Keilwagen J. PRROC: computing and visualizing precision-recall and receiver operating characteristic curves in R. Bioinformatics. 2015;31(15):2595–2597.

70. Smyth GK. limma: Linear Models for Microarray Data. In: Gentleman R, Carey VJ, Huber W, Irizarry RA, Dudoit S, eds. Bioinformatics and Computational Biology Solutions Using R and Bioconductor. Springer New York; 2005:397–420.

71. Supek F, Bošnjak M, Škunca N, Šmuc T. REVIGO summarizes and visualizes long lists of gene ontology terms. PLoS One. 2011;6(7):e21800.

72. Su G, Morris JH, Demchak B, Bader GD. Biological network exploration with Cytoscape 3. Curr Protoc Bioinformatics. 2014;47:8.13.1-24.

