## Supplemental Materials for "A Highly Accurate Ensemble Classifier for the Molecular Diagnosis of ASD at Ages 1 to 4 Years"

   Vahid H. Gazestani, Bokan Bao, Yaqiong Xiao, Srinivasa Nalabolu, Karen Pierce & Eric Courchesne
2. Department of Pediatrics, University of California San Diego, La Jolla, CA, USA

   Vahid H. Gazestani, Bokan Bao, Austin W.T. Chiang & Nathan E. Lewis
3. Bioinformatics and Systems Biology Program, University of California San Diego, La Jolla, CA, USA

   Bokan Bao & Nathan E. Lewis
4. Department of Bioengineering, University of California San Diego, La Jolla, CA, USA

   Bokan Bao & Nathan E. Lewis
5. Department of Computer Science, University of North Carolina, Chapel Hill, NC, USA

   Raphael Kim
6. Renaissance Computing Institute, The University of North Carolina at Chapel Hill, Chapel Hill, NC, USA

   Raphael Kim & Kimberly Robasky

7. Department of Genetics, University of North Carolina at Chapel Hill, Chapel Hill, NC 27514, United States
   Kimberly Robasky
8. School of Information and Library Science, University of North Carolina at Chapel Hill, Chapel Hill, NC 27599, United States
   Kimberly Robasky
9. Carolina Health and Informatics Program, University of North Carolina at Chapel Hill, Chapel Hill, NC 27599, United States
   Kimberly Robasky

*These first authors contributed equally: Vahid H. Gazestani and Bokan Bao

‡ Corresponding co-equal Senior authors are Nathan E. Lewis and Eric Courchesne

Names: Nathan E. Lewis

Address: 9500 Gilman Drive MC 0760, La Jolla, CA 92093

Name: Eric Courchesne

Address: 8110 La Jolla Shores Dr #201, La Jolla, CA 92037

**Content**

**eResult 1** Sanity check for 1822 high-performance model on Longitudinal dataset

**eResult 2** Validation of Independent Replication dataset

**eResult 3** Result and parameter setting on the baseline Random Forest model

**Supplemental Figure 1**

**Supplemental Figure 2**

**Supplemental Figure 3**

**Supplemental Figure 4**

**Supplemental Figure 5**

**Supplemental Figure 6**

**Supplemental Figure 7**

**Supplemental Figure 8**

**eResult 1**

**Sanity check for 1822 high-performance model.**

First, we evaluated the performance of the 1822 high-performing models on a Longitudinal dataset. of 18 ASD and 15 TD samples from toddlers who were included in the Discovery dataset. Out of 1822 models with AUC-ROC of above 0.8, 1656 (90.8%, odds ratio 76.89514, 95% confidence interval [64.74134, 91.39221], two-sided Fisher's Exact Test $P < 2.2e-16$) of the models showed AUC-ROC above 0.8 in the Longitudinal dataset.

**eResult 2**

**Validation of Independent Replication dataset**

To test the sensitivity, the models were trained on the Discovery dataset (175 subjects together) and then tested on the dataset with 34 ASD and 31 TD. The AUC-ROC values were used to evaluate the performance of the selected 1822 high-performing models. 1076 models (59.0%) had an AUC-ROC value greater than 0.75. The two-sided Fisher's Exact Test calculated the odds ratio 5.133581 (95% confidence interval [4.688261, 5.620152]) with $P < 2.2e-16$.

In terms of the diagnostic specificity, the models were trained on the discovery dataset (175 subjects together) and then tested on the independent Replication dataset (55 subjects, 31 TD, and 24 LD). From 1822 models with AUC-ROC or AUC-PR values greater than 0.8, none had an AUC-ROC value of above 0.8 in this LD vs TD dataset. The two-sided Fisher's Exact Test calculated the odds ratio 0 (95% confidence interval [0, 0.01765617]) with $P < 2.2e-16$. 26 methods had an AUC-ROC value of above 0.8 in this LD vs TD dataset. The two-sided Fisher's Exact Test calculated the odds ratio 0.1240666 (95% confidence interval [0.08055236, 0.18305634]) with $P < 2.2e-16$.

We also assessed the diagnostic specificity of the models by comparing performance of the 1822 models in separating a Diagnostic Specificity dataset of 24 language delayed (LD) toddlers from the ASD samples. From 1822 models with AUC-ROC or AUC-PR above 0.8, 26 (1.42%; odds ratio 0.1113861, 95% confidence interval [0.07231532, 0.16436198]), two-sided Fisher's Exact Test $P < 2.2\text{e-}16$) have an AUC-ROC above 0.75 in this LD vs TD dataset.

**eResult 3**

**Result and parameter setting on the baseline Random Forest model**

Number of top variables are chosen from results based on the validation data, over a grid of (10, 100, 500, 1000). On this grid, 500 genes are chosen as optimal. Importances are generated from 100 rounds of evaluations of Random Forest, averaged. Final test results using these top 500 genes have the accuracy: 73.33%, sensitivity: 85.71%, specificity: 62.50%, AUC-ROC: 72.32%.

**Supplementary Figure 1. The detail workflow of the feature engineering pipeline**

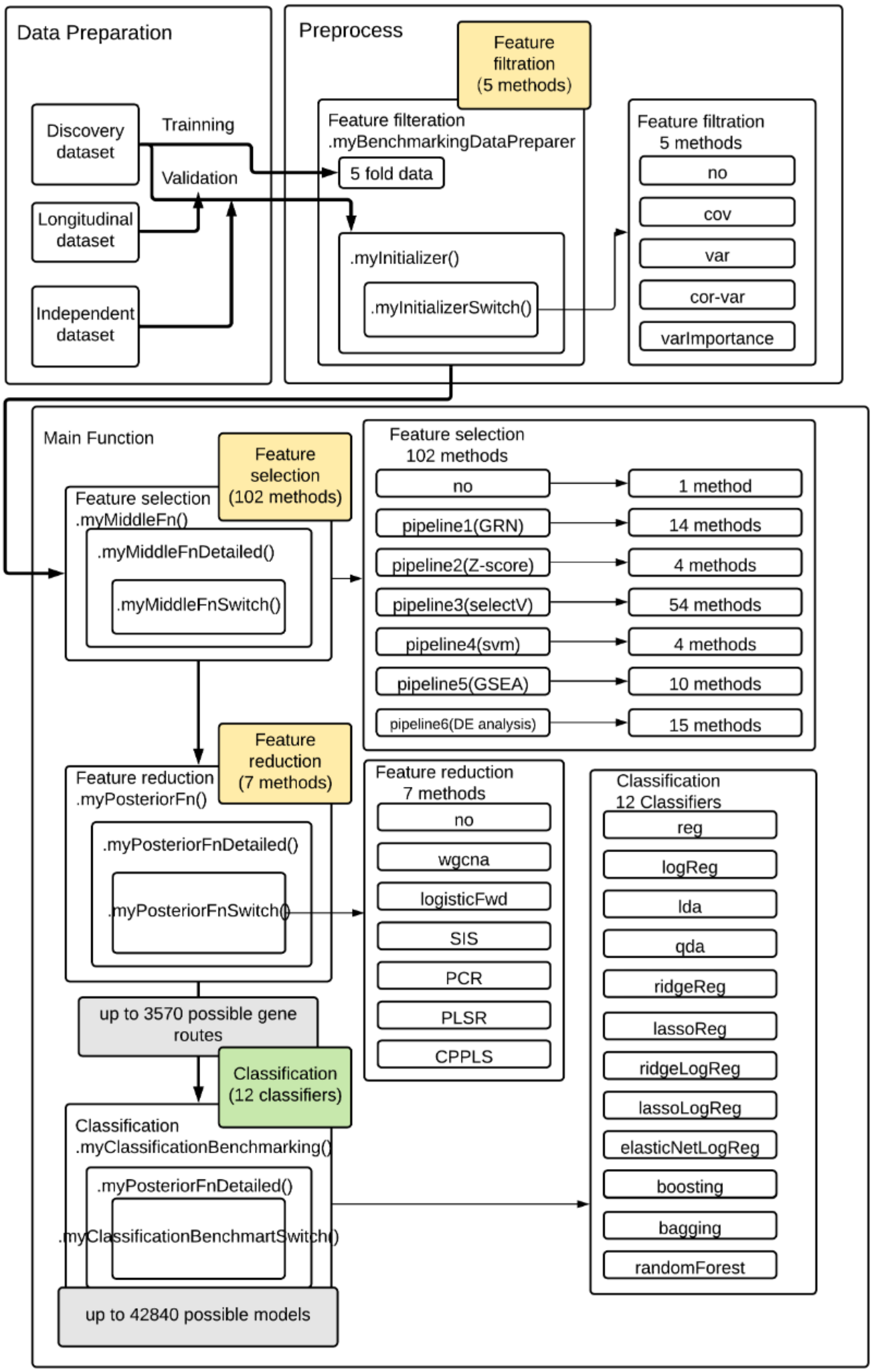

**Supplementary Figure 2. The overall data processing workflow** The workflow lists the processes of running main training test on Discovery dataset, validation test on independent Replication dataset, reproducibility test on longitudinal dataset and background test on randomized permutation dataset.

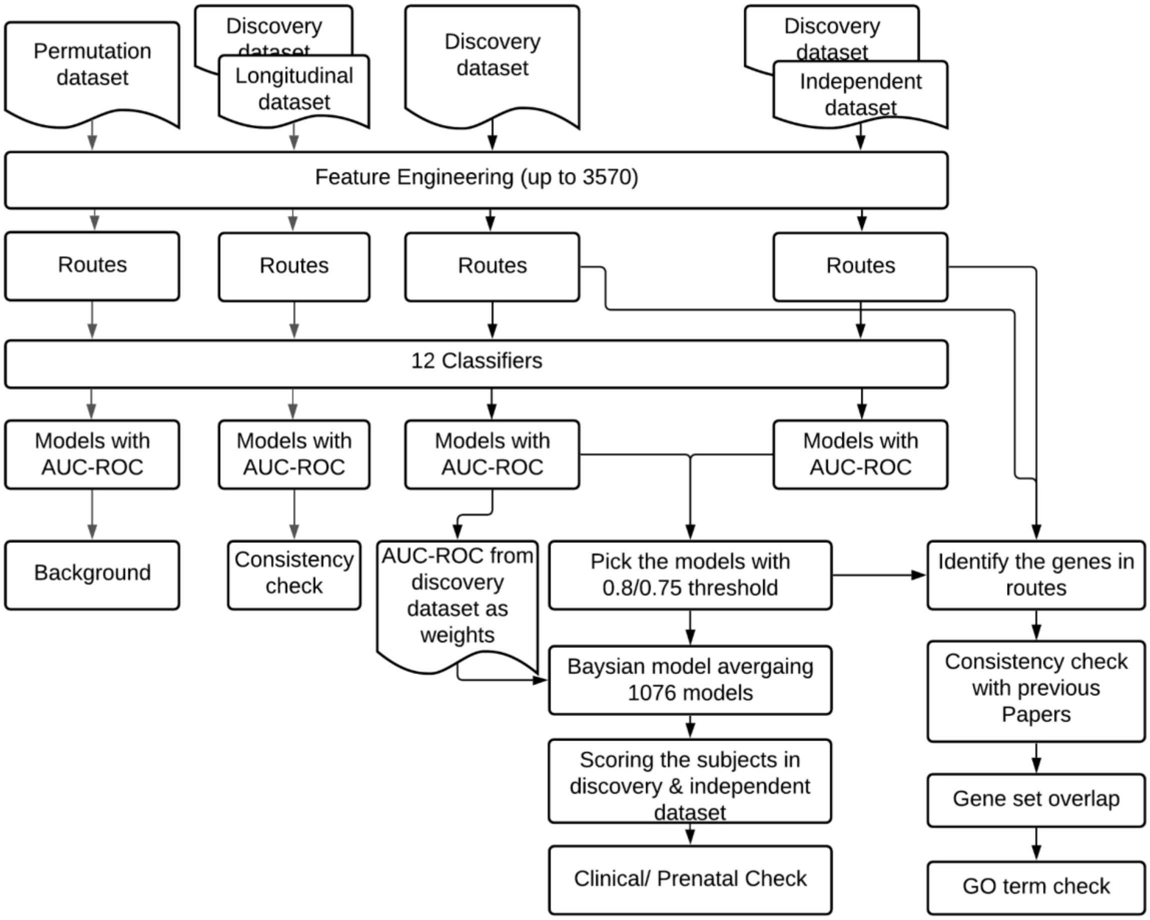

**Supplementary Figure 3. The distribution of AUC-ROC scores of 12 classifiers against the AUC-ROC mean score of 1320 routes.** The distribution of AUC-ROC generated by 1320 routes within each of 12 classifiers in **A.** the discovery dataset and **B.** the independent dataset. The distribution of AUC-ROC generated by 1320 routes vs the mean of AUC-ROC of each route in **C.** the discovery dataset (with +- 0.0514 95% confidence interval) and **D.** the distribution in the independent dataset (with +- 0.0514 95% confidence interval) . The variance of AUC-ROC generated by 12 classifiers of 1320 routes, the x-axis is the mean of AUC-ROC of each route in **E.** the main dataset and **F.** in the test dataset.

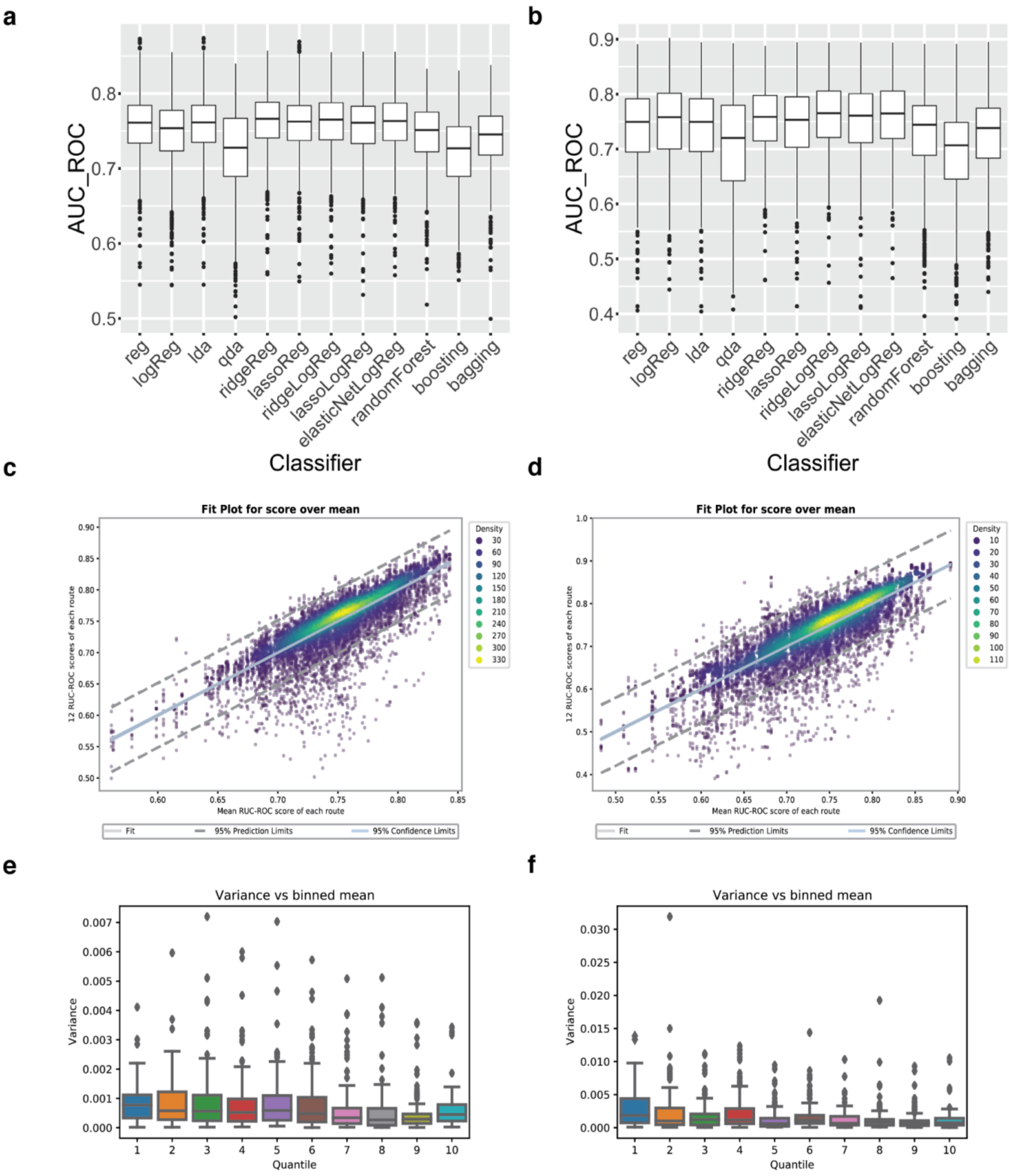

a
AUC_ROC
0.5
0.6
0.7
0.8
reg
logReg
lda
qda
ridgeReg
lassoReg
ridgeLogReg
lassoLogReg
elasticNetLogReg
randomForest
boosting
bagging
Classifier
b
AUC_ROC
0.4
0.5
0.6
0.7
0.8
0.9
reg
logReg
lda
qda
ridgeReg
lassoReg
ridgeLogReg
lassoLogReg
elasticNetLogReg
randomForest
boosting
bagging
Classifier
c
Fit Plot for score over mean
12 RUC-ROC scores of each route
Mean RUC-ROC score of each route
Density
30
60
90
120
150
180
210
240
270
300
330
Fit
95% Prediction Limits
95% Confidence Limits
d
Fit Plot for score over mean
12 RUC-ROC scores of each route
Mean RUC-ROC score of each route
Density
10
20
30
40
50
60
70
80
90
100
110
Fit
95% Prediction Limits
95% Confidence Limits
e
Variance vs binned mean
Variance
Quantile
f
Variance vs binned mean
Variance
Quantile

**Supplementary Figure 4. The AUC-ROC score of 1320 models trained on Discovery dataset. A.** The score distribution of 1822 models that have AUC-ROC score above 0.8 in the discovery dataset in 5 iterations. **B.** The score-difference distribution of 1822 models in 5 iterations.

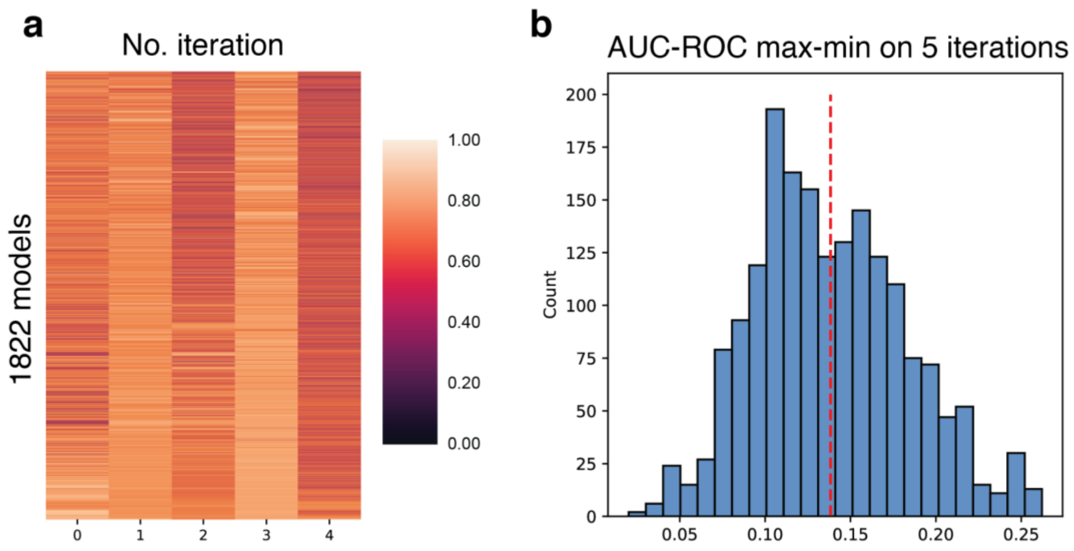

**Supplementary Figure 5**

**The geneset similarity distance among 191 routes.** The distance score (see **Methods**) is measured by the top 500 genes that are used by 191 routes and then clustered by 'average' method. The data of the table is in **supplementary table 5**.

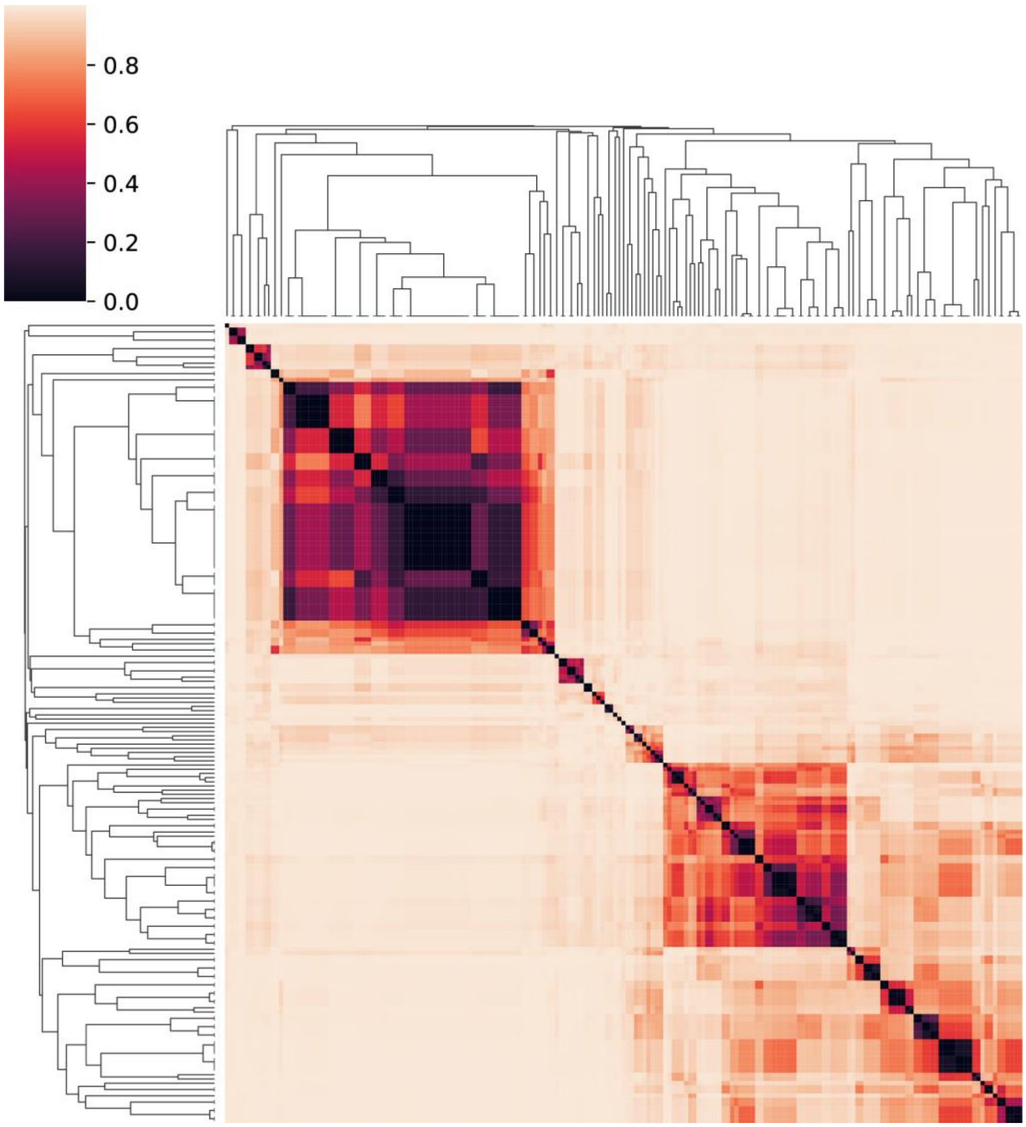

**Supplementary Figure 6. The relation between the similarity of gene routes and the 12 classifiers behavior.**

The correlation of 12-classifier score vs the distance between routes and models. The distance is measured by the -log2 of the geneset similarity on the first 500 genes (see **Methods**). The slope=-0.03806252570865483, intercept=0.4443827187728834, rvalue=-0.25294548575900266, pvalue=1.35416223411553e-265, stderr=0.001075187710703305

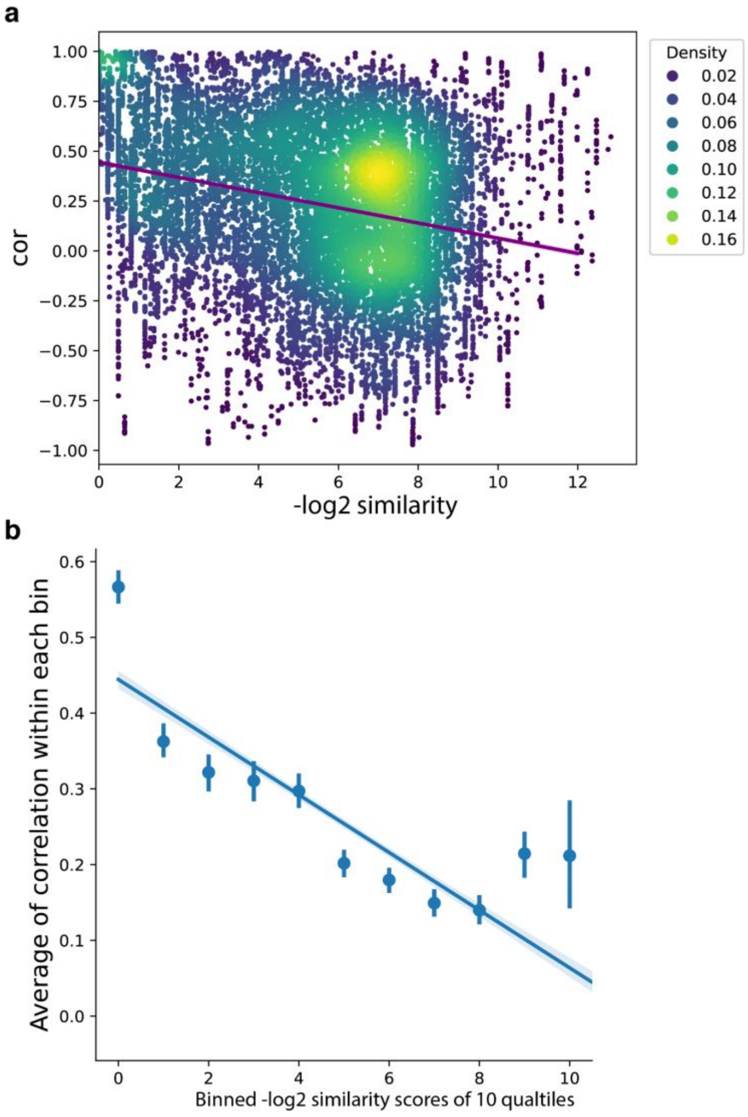

**Supplementary Figure 7. Illustration of the updated model that combines the composite model with geo-fixation model.** The updated model was tested on 132 of 175 Discovery dataset subjects and 41of 65 Replication dataset subjects who had available Geo-Fixation data (e.g., moderate or good data quality, total looking time > 50%). By directly classifying the subjects who had percent fixation on non-social images >69% as ASD (GeoPref-subtype). **A**. The composite score vs geo-fixation percentage score in discovery dataset. **B**. The updated score vs geo-fixation percentage score for discovery dataset. **C**. The composite score vs geo-fixation percentage score for the independent dataset. **D**. The updated score vs geo-fixation percentage score for independent dataset.

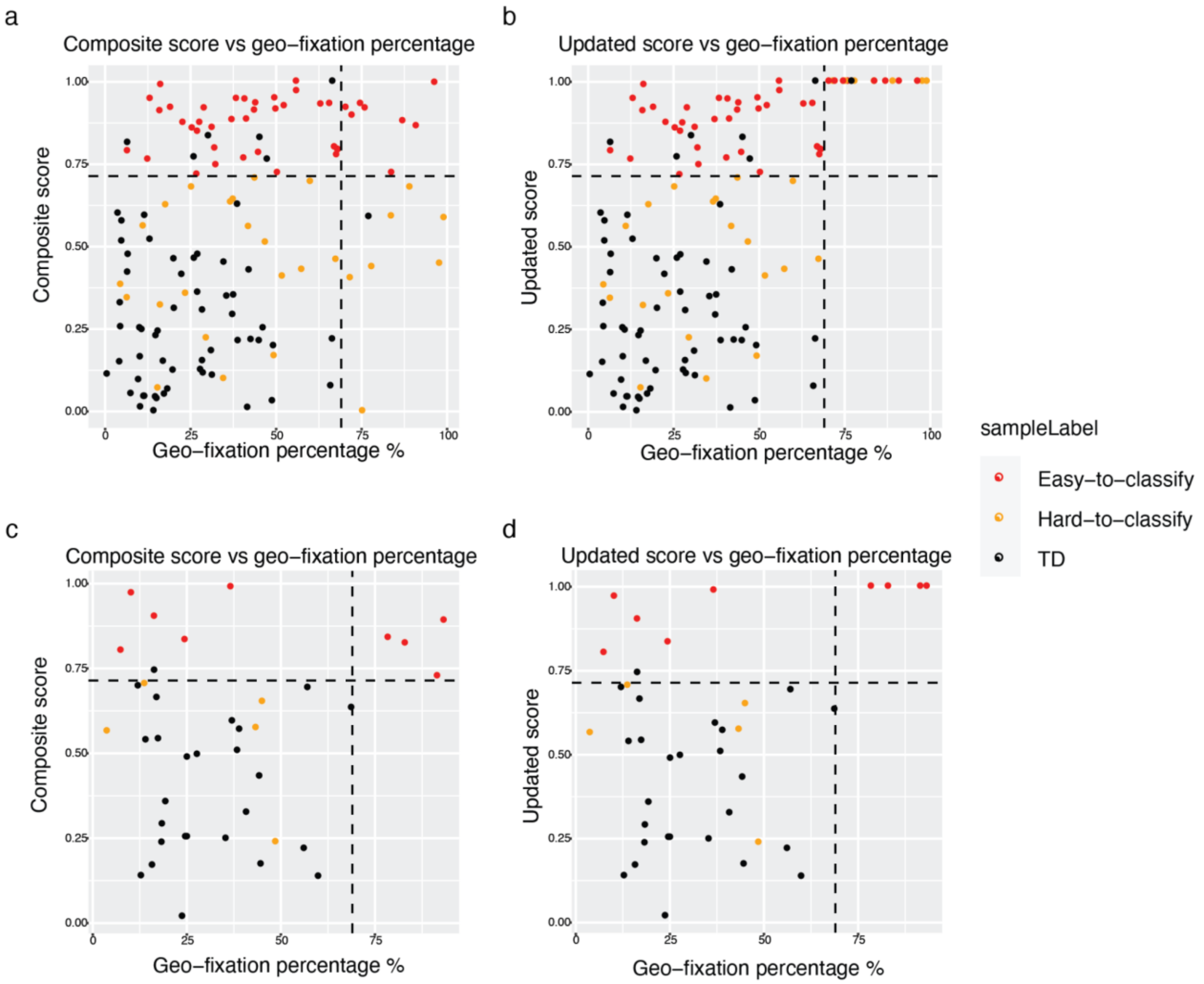

**Supplementary Figure 8.** Diagnostic and psychometric scores were not significantly different between ASD toddlers above (easy-to-classify) and below the mean composite classifier score (hard-to-classify). **A-B** discovery dataset r= 0.5310815, Two sided t.test P = 4.011e-14 **C-D** independent dataset r= 0.4871145, Two sided t.test P = 3.873e-05

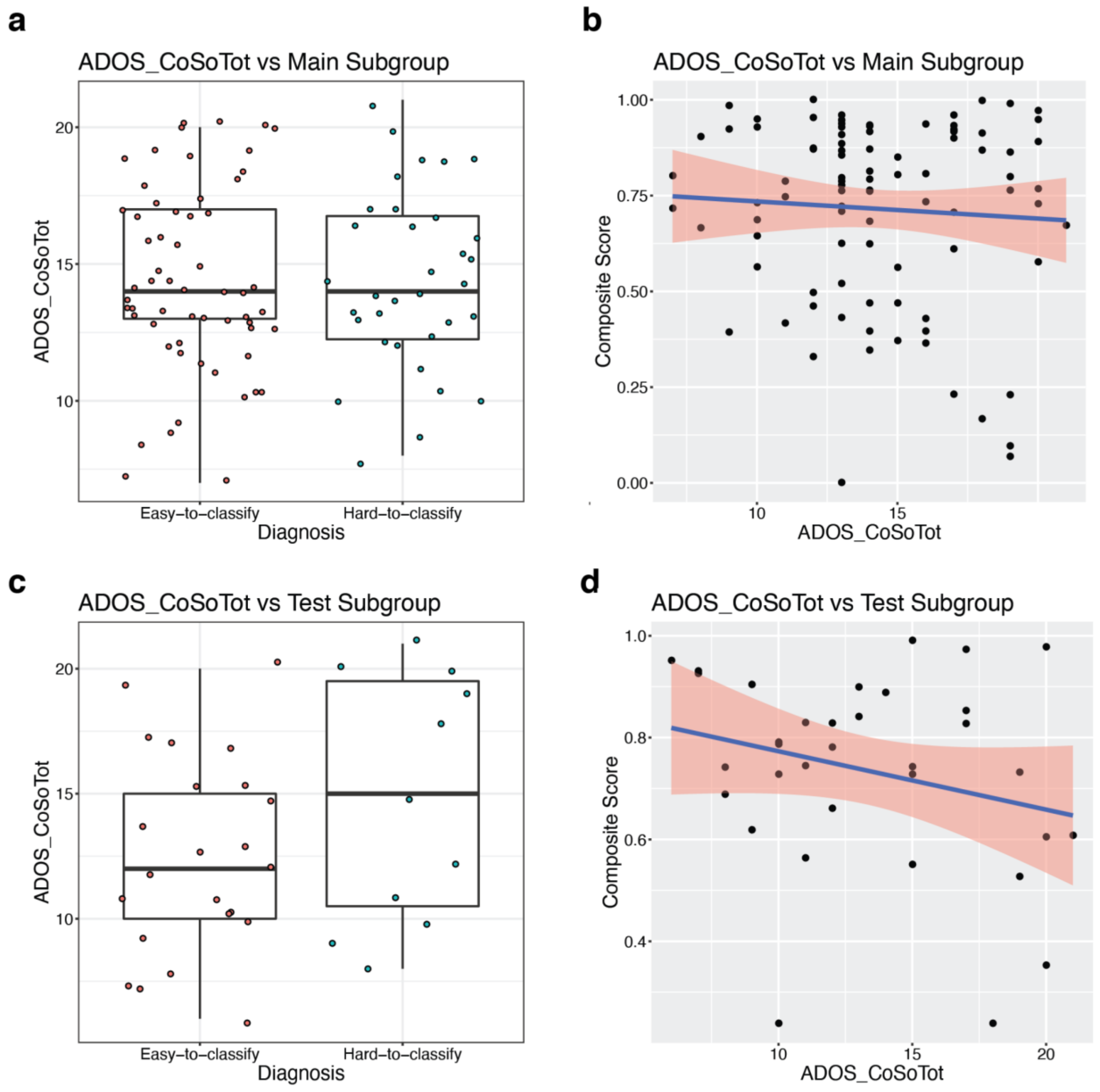
